# Impact of Adverse Drug Reactions in the outcomes of Tuberculosis Treatment

**DOI:** 10.1101/2022.05.31.22275820

**Authors:** Flávia M. Sant’Anna, Mariana Araújo-Pereira, Carolina A. S. Schmaltz, María B. Arriaga, Bruno B. Andrade, Valeria C. Rolla

## Abstract

Adverse drug reactions (ADR) challenge successful anti-tuberculosis treatment (ATT). The aim of this study was to evaluate the impact of ATT-associated ADR and related factors on treatment outcomes. A prospective cohort study of persons with tuberculosis at a referral center in Rio de Janeiro, Brazil, from 2010 to 2016. ADRs, favorable (cure and treatment completion) and unfavorable (death, loss to follow up and failure) outcomes were prospectively captured. The Kaplan-Meier curve was used to estimate probability ADR-free time. A logistic regression model was performed to identify independent associations with unfavorable outcomes. 550 patients were enrolled and 35.1% were people living with HIV (PLHIV). ADR occurred in 78.6% of participants and was associated with favorable outcomes. Smokers (OR:2.32;95%CI:1.34-3.99) and illicit drug users (OR:2.02;95%CI:1.15-3.55) exhibited higher risk of unfavorable outcome. PLHIV more frequently experienced grade 3/4-ADR, specially “liver and biliary system disorders”. Lower CD4 counts were associated with hepatotoxicity (p=0.03). Male sex, low schooling, smoking and illicit-drug use are independent risk factors for unfavorable outcomes, as well alcohol abuse and previous ART in PLHIV. ADR increases the odds of favorable outcomes, although it is more severe in PLHIV and related to a higher risk of hepatotoxicity.

## Introduction

Anti-tuberculosis treatment (ATT) has been available worldwide and can reach more than 90% of effectiveness [1]. However, the challenges in achieving this goal persist as ATT can cause adverse drug reactions (ADR) that lead to increased morbidity and compromise adherence eventually contributing to treatment failure, relapse, or emergence of resistant strains [2,3].

The ATT recommended by World Health Organization (WHO) and implemented since 2009 in the Brazilian guidelines is as a fixed-dose (FDC), single-tablet combination of four drugs (rifampicin, isoniazid, pyrazinamide, and ethambutol) during the intensive phase of treatment followed by rifampicin and isoniazid in FDC until the end of treatment [4]

Retrospective studies conducted in Brazil found an incidence of ADR between 23% to 83% [5– 7] and in a meta-analysis including other non-Brazilian studies, the incidence varied from 8.4% to 83.5% [8]. ADRs are multifactorial [9,10] and the major determinants are unadjusted prescribed doses of medications, patient’s age, nutritional status, alcohol consumption, liver and kidney function and HIV-infection [11,12].

In people living with HIV (PLHIV), co-administration of antiretroviral therapy (ART) and ATT increases the risk of drug interactions, immunopathological responses, and ADRs [13]. ADRs have been associated with almost two-fold increased risk of unsuccessful treatment outcomes after 6 months of ATT [14].

According to data from the Brazilian Ministry of Health, in the year of 2020, 8.5% of new diagnoses of tuberculosis were in PLHIV [15]. ART during ATT is challenging because of drug-to-drug interactions and increased risk of ADR. In Brazil there are few studies on ADR in PLHIV treating TB, with no definitive conclusion. The aim of this study is to evaluate the impact of ADR due to ATT and factors associated with favorable and unfavorable outcomes. We hypothesize that ADR occurrence could increase unfavorable outcome occurrences.

## Methods

### Ethics statement

The study was approved by the Institutional Review Board of the National Institute of Infectious Diseases Evandro Chagas (INI) (CAAE: 86215118.5.0000.5262). Written informed consent was obtained from all participants involved in the study and all clinical investigations were conducted according to the principles expressed in the Declaration of Helsinki.

### Study design

We conducted a prospective cohort study with TB patients treated and followed up at the Clinical Research Laboratory on Mycobacteria (LAPCLIN-TB) of the National Institute of Infectious diseases Evandro Chagas (INI), Oswaldo Cruz Foundation (FIOCRUZ), Rio de Janeiro, Brazil, from January 2010 to December 2016. In LAPCLIN-TB a cohort study has been active since 2000, with data registered at a standardized visit template. The information is captured at each patient’s visit. Raw data is shown in S1 File.

### Inclusion and exclusion criteria

The inclusion criteria were 18 years old or more and pulmonary, extrapulmonary or disseminated TB. The exclusion criteria were death or loss to follow up (LTFU) within the first 15 days of ATT (early death or early LTFU avoiding data capture), rifampicin and isoniazid resistance (MDR) and lack of information on outcomes. Patients initially diagnosed with TB based on clinical-radiologic evaluation who were later diagnosed with another disease rather than tuberculosis were also excluded.

### TB diagnosis and follow up visits

Tuberculosis diagnosis was clinical-radiological (based on symptoms and signs and chest X-ray findings), in some cases confirmed by laboratory tests (acid fast smears and/or culture and/or histopathological findings and/or Xpert-MTB/RIF™). In cases without laboratory confirmation, a positive therapeutic response to ATT was considered for TB diagnosis (clinical-radiological improvement with ATT). Visits were scheduled at baseline (ATT initiation), 15, 30, 60, 90, 120, 150 and 180 days after ATT initiation. Sometimes treatment was prolonged, especially for disseminated or extrapulmonary TB cases and due to a physician decision. In these cases, there were monthly visits scheduled after the 180 days of ATT. Data were entered into an electronic medical record and standardized information was collected using a predefined template in all visits.

During the baseline visit, information collected included demographic variables such as age, sex, race (self-reported), marital status, schooling (illiterate, elementary school up to 12 years, and high schooling above 12 years), social behavior (alcohol, tobacco, illicit drugs use/addiction), clinical information such as clinical forms of tuberculosis (pulmonary, extrapulmonary or disseminated - 2 or more non-contiguous sites), presence of comorbidities, and concomitant medications use. At baseline visits laboratory tests were collected: hemogram, urea, creatinine, uric acid, alanine (ALT) and aspartate (AST) transferases, albumin, alkaline phosphatase, gamma glutamyl transferase (GGT), chest X-ray, hepatitis B, C and HIV serology. At subsequent visits, ADR were monitored with clinical information and laboratory evaluation (hemogram and biochemistry including liver enzymes) in all visits. For known PLHIV, CD4 count, and HIV viral load (VL) were added at baseline visits and at the end of ATT.

### Anti-TB treatment (ATT)

Since 2009, Brazilian guidelines incorporated WHO-recommendation of rifampicin 600 mg, isoniazid 300 mg, pyrazinamide 1600 mg and ethambutol 1100 mg in FDC for two months, followed by four months of rifampicin 600 mg and isoniazid 300 mg in FDC, for both new and retreatment TB cases until drug-susceptibility tests were available. Doses were adjusted for patients with less than 50kg [4].

### HIV treatment

ART was prescribed for PLHIV, by each assistant physician, according to Brazilian guidelines for Sexual Transmitted Diseases and AIDS [16]. The recommended ATT for PLHIV included a rifamycin (rifampicin or rifabutin), which was chosen according to the ART regimen appropriate for each specific patient. Rifabutin 150 mg/day was chosen when protease inhibitors were prescribed as part of ART [11]. ART was introduced for naïve patients as soon as possible, which happened around the first month of ATT in most cases.

### Smoke habit

A current smoker was defined as someone who was smoking at the time of tuberculosis diagnosis and/or at the beginning of ATT or who stopped smoking due to tuberculosis symptoms but went back to smoke during ATT.

### Illicit drug users

All patients who reported to be currently using drugs were considered illicit drug users.

### Alcohol abusers

All patients answered the CAGE questionnaire [17] and those who answered yes to 2 or more questions were considered as having significant alcohol addiction.

### Adverse Drug Reaction (ADR)

ADR were defined according to signs and symptoms (clinical), laboratory tests abnormalities (laboratory) or both (clinical and laboratory). ADRs were categorized according to WHO-ART classification [18]. The scale of intensity was based on the Division of AIDS table for grading the severity of adult and pediatric ADR [19]: Grade 1: mild event, Grade 2: moderate event, Grade 3: severe event, Grade 4: potentially life-threatening event, Grade 5: death. The causality of ADRs were assessed using the Naranjo Scale, as improbable, possible, probable, and definitively related to the medications [20]. Only probable, possible, and definitively related ADR were considered for the analysis.

### Outcomes

Outcomes were based on WHO criteria [21]. The success of treatment consisted of cure and treatment conclusion which were considered favorable outcomes. Unfavorable outcomes were grouped (composite outcome) and included treatment failure (remaining smear-positive after the 4th month of ATT), transfer to different treatment facilities (patients who were transferred out to other health unit), LTFU (interruption of treatment for two or more consecutive months) and death associated with tuberculosis (all cases were revised and the causality accessed).

## Data analyses

Descriptive statistics was performed using the median values with interquartile ranges (IQR) as measures of central tendency and dispersion for continuous variables. Categorical variables were described using frequency (no.) and proportions (%). The Mann–Whitney U test (for two unmatched groups) was used to compare continuous variables and the Pearson chi-square test was used to compare categorical variables between study groups. The Kaplan-Meier curve was used to estimate ADR-free probabilities.

A logistic binary regression was performed with backward stepwise selection using variables with univariate p-value ≤0.2 to identify independent associations between characteristics of tuberculosis patients with the composite unfavorable treatment outcome. Results from regression were presented in terms of point estimates and 95% confidence intervals (CI). All analyses were pre-specified. Differences with p-values below 0.05 after adjustment for multiple comparisons (Holm-Bonferroni) were considered statistically significant.

The statistical analyses and data visualization were performed using rstatix (version 0.4.0), stats (version 3.6.2), ggplot2 (version 3.3.2), survivor (version 3.2.7) and survminer (version 0.4.8) R packages.

## Results

From 2010 to 2016, 606 patients were enrolled in this study. Fifty-six patients with exclusion criteria were ruled out: 31 with MDR tuberculosis, 18 with other diagnoses during ATT (including non-tuberculous mycobacteria), two due to early death, and one because of early LTFU. Two patients were removed from the dataset due to lack of information on the outcomes.

A total of 550 patients were included in this study. Only patients with ADR (total 426) were included in these analyses **(Figure 1A). Table 1** shows a comparison of favorable and unfavorable outcomes regarding their sociodemographic and clinical characteristics. The median age was 38 years, 329 (59.8%) were male and 193 (35.1%) were PLHIV. Among patients with unfavorable outcomes: 84 were LTFU, 7 had a treatment failure and 7 died.

**Table 1.** Clinical, social demographics and comorbidities according to outcomes.

|  | **All (n=550)** | **Favorable**<br>**(n=452)** | **Unfavorable**<br>**(n=98)** | **p**<br>**value** |
| --- | --- | --- | --- | --- |
| **Male, n (%):** | 329 (59.8) | 256 (56.6) | 73 (74.5) | **0.002** |
| **Age, median (IQR):** | 38 (29-49) | 39 (29-50) | 36 (28.2-46.8) | 0.138 |
| **Race, n (%):** |  |  |  | **<0.001** |
| White | 281 (51.1) | 250 (55.3) | 31 (31.6) |  |
| Black/Pardo | 269 (48.9) | 202 (44.7) | 67 (68.4) |  |
| **Scholarity, n (%):** |  |  |  | **<0.001** |
| Illiterate | 21 (3.82) | 16 (3.54) | 5 (5.10) |  |
| Elementary<br>School | 449 (81.6) | 356 (78.8) | 93 (94.9) |  |
| High School | 80 (14.5) | 80 (17.7) | 0 (0.00) |  |
| **Married, n (%):** | 279 (50.7) | 239 (52.9) | 40 (40.8) | **0.040** |
| **Tobacco use, n (%):** | 191 (34.7) | 134 (29.6) | 57 (58.2) | **<0.001** |
| **Alcohol abuse, n (%):** | 212 (38.5) | 156 (34.5) | 56 (57.1) | **<0.001** |
| **Illicit drug use, n (%):** | 121 (22.0) | 78 (17.3) | 43 (43.9) | **<0.001** |
| **Initial Weight, median (IQR):** | 58 (50.6-67.5) | 59.7 (50.7-9.6) | 56.3 (50.1-64) | **0.023** |
| **ADR occurrence, n (%):** | 431 (78.6) | 372 (82.5) | 59 (60.8) | **<0.001** |
| **Type of TB, n (%):** |  |  |  | **0.023** |
| PTB | 290 (52.7) | 238 (52.7) | 52 (53.1) | 1 |
| EPTB | 170 (30.9) | 148 (32.7) | 22 (22.4) | 0.060 |
| Disseminated | 90 (16.4) | 66 (14.6) | 24 (24.5) | **0.025** |
| **Changed treatment, n (%):** | 64 (11.6) | 49 (10.8) | 15 (15.3) | 0.282 |
| **Hepatitis B or C, n (%):** | 28 (5.09) | 23 (5.09) | 5 (5.10) | 1 |
| **Diabetes mellitus, n (%)**: | 50 (9.09) | 46 (10.2) | 4 (4.08) | 0.087 |
| **Hypertension, n (%)**: | 78 (14.2) | 72 (15.9) | 6 (6.12) | **0.018** |
| **COPD, n (%)**: | 10 (1.82) | 9 (1.99) | 1 (1.02) | 1 |
| **HIV infection, n (%)**: | 193 (35.1) | 141 (31.2) | 52 (53.1) | **<0.001** |
| ART naive, n (%) | 74 (38.7) | 62 (44.3) | 12 (23.5) | **0.015** |

**Figure 1.**
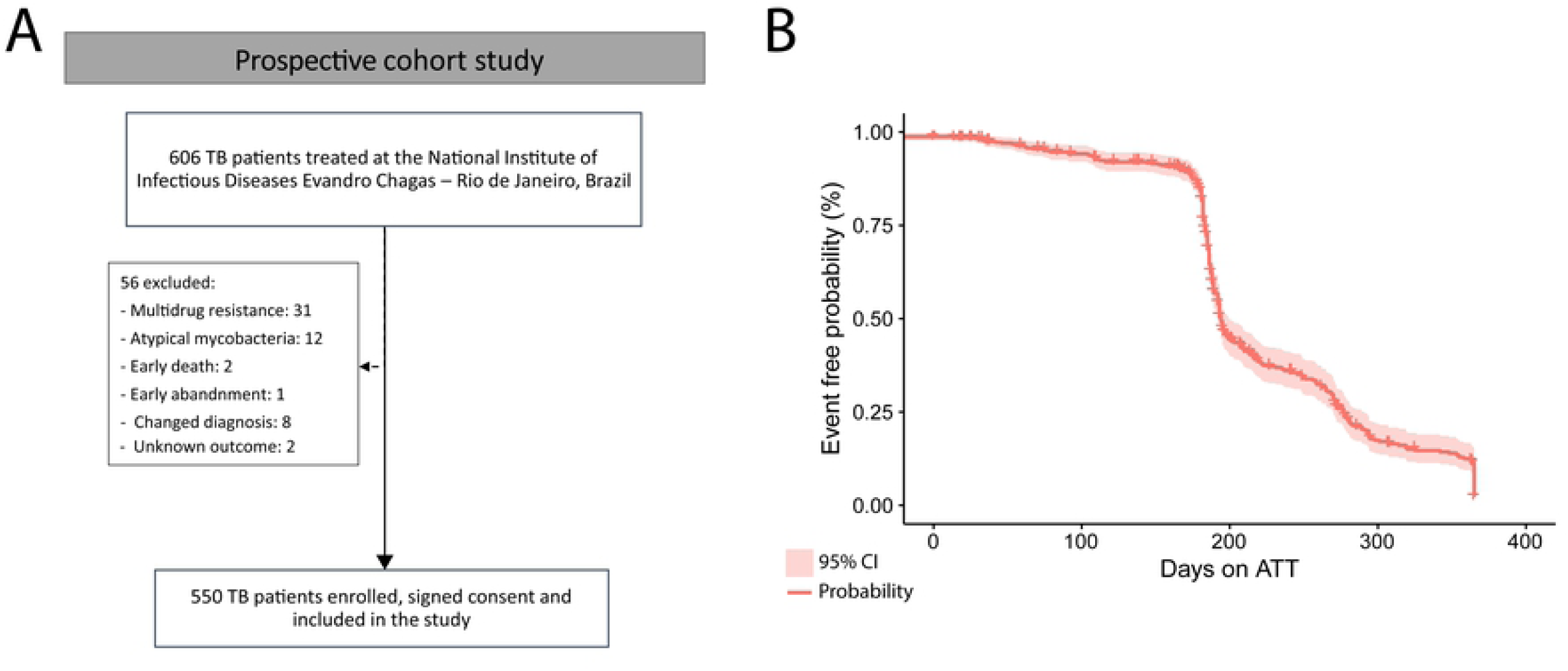
Study Design and probability of presenting adverse reactions during the antitubercular treatment. (A) Study design. (B) Event free curve (Kaplan-Meier) of Tuberculosis patients during antitubercular treatment. Mean time (in state, restricted max time = 365) in days of event occurrence: 140.7. Abbreviation: ADR: adverse drug reaction; ATT: anti-TB treatment; CI: confidence interval; TB: tuberculosis.

All participants who died were PLHIV with clinical and/or laboratorial signs of advanced immunodeficiency. No deaths were related to ADR. We observed significant differences in race, schooling, smoking habits, and alcohol consumption among cases with unfavorable outcomes when compared to patients who experienced favorable outcomes. ADR occurred in 78.6% of participants, 35.1% of ADR were in PLHIV. Despite the high incidence, occurrence of ADR was higher in the group of participants who experienced favorable ATT outcomes.

Our analysis of the type, severity, date of onset, relationship with the ATT and WHO-ART classification according to outcomes are presented in **Table 2**. According to WHO-ART classification, gastro-intestinal system disorders (23.9%) followed by “metabolic and nutritional disorders” (22.4%), were the most prevalent ADR related to unfavorable outcomes. The probability of ADR was higher until 140 days of ATT **(Figure 1B)**. In the logistic regression model, smoking habit, illicit drugs use, and HIV-infection were associated with unfavorable outcomes **(Figure 2)**.

**Table 2.** ADR characteristics according to ATT outcome.

|  | All (n=695) | Favorable<br>(n=628) | Unfavorable<br>(n=67) | p value |
| --- | --- | --- | --- | --- |
| **Type of event, n (%)** |  |  |  | **0.95** |
| **Clinical + Laboratory** | **87 (12.5)** | **78 (12.4)** | **9 (13.4)** |  |
| **Laboratory** | **228 (32.8)** | **207 (33.0)** | **21 (31.3)** |  |
| **Clinical** | **380 (54.7)** | **343 (54.6)** | **37 (55.2)** |  |
| **Adverse reactions duration (days), median (IQR)** | **77.0 (39.0-142)** | **81.5 (43.0-147)** | **44.0 (25.0-85.0)** | **<0.001** |
| **Severity of ADR, n (%)** |  |  |  | **0.128** |
| **Grade 1** | **264 (38.0)** | **246 (39.2)** | **18 (26.9)** |  |
| **Grade 2** | **352 (50.6)** | **314 (50.0)** | **38 (56.7)** |  |
| **Grade 3** | **67 (9.64)** | **57 (9.08)** | **10 (14.9)** |  |
| **Grade 4** | **12 (1.73)** | **11 (1.75)** | **1 (1.49)** |  |
| **Onset of ADR, n (%)**: |  |  |  | **0.38** |
| **before 2nd month (early)** | **592 (85.2)** | **532 (84.7)** | **60 (89.6)** |  |
| **after month 2 (late)** | **103 (14.8)** | **96 (15.3)** | **7 (10.4)** |  |
| **Relation of ADR with ATT, n (%)**: |  |  |  | **0.601** |
| **Related** | **14 (2.01)** | **14 (2.23)** | **0 (0.00)** |  |
| **Probable** | **171 (24.6)** | **153 (24.4)** | **18 (26.9)** |  |
| **Possible** | **510 (73.4)** | **461 (73.4)** | **49 (73.1)** |  |
| **WHO ART classification, n (%)**: |  |  |  | **0.207** |
| **Central and peripheral nervous system disorders** | **45 (6.47)** | **43 (6.85)** | **2 (2.99)** |  |
| **Gastro-intestinal system disorder** | **151 (21.7)** | **135 (21.5)** | **16 (23.9)** |  |
| **Liver and biliary system disorders** | **54 (7.77)** | **44 (7.01)** | **10 (14.9)** |  |
| **Metabolic and nutritional disorders** | **171 (24.6)** | **156 (24.8)** | **15 (22.4)** |  |
| **Musculo-skeletal disorders** | **63 (9.06)** | **56 (8.92)** | **7 (10.4)** |  |
| **Other** | **46 (6.62)** | **45 (7.17)** | **1 (1.49)** |  |
| **Skin and appendages disorders** | **125 (18.0)** | **113 (18.0)** | **12 (17.9)** |  |

**Figure 2.**
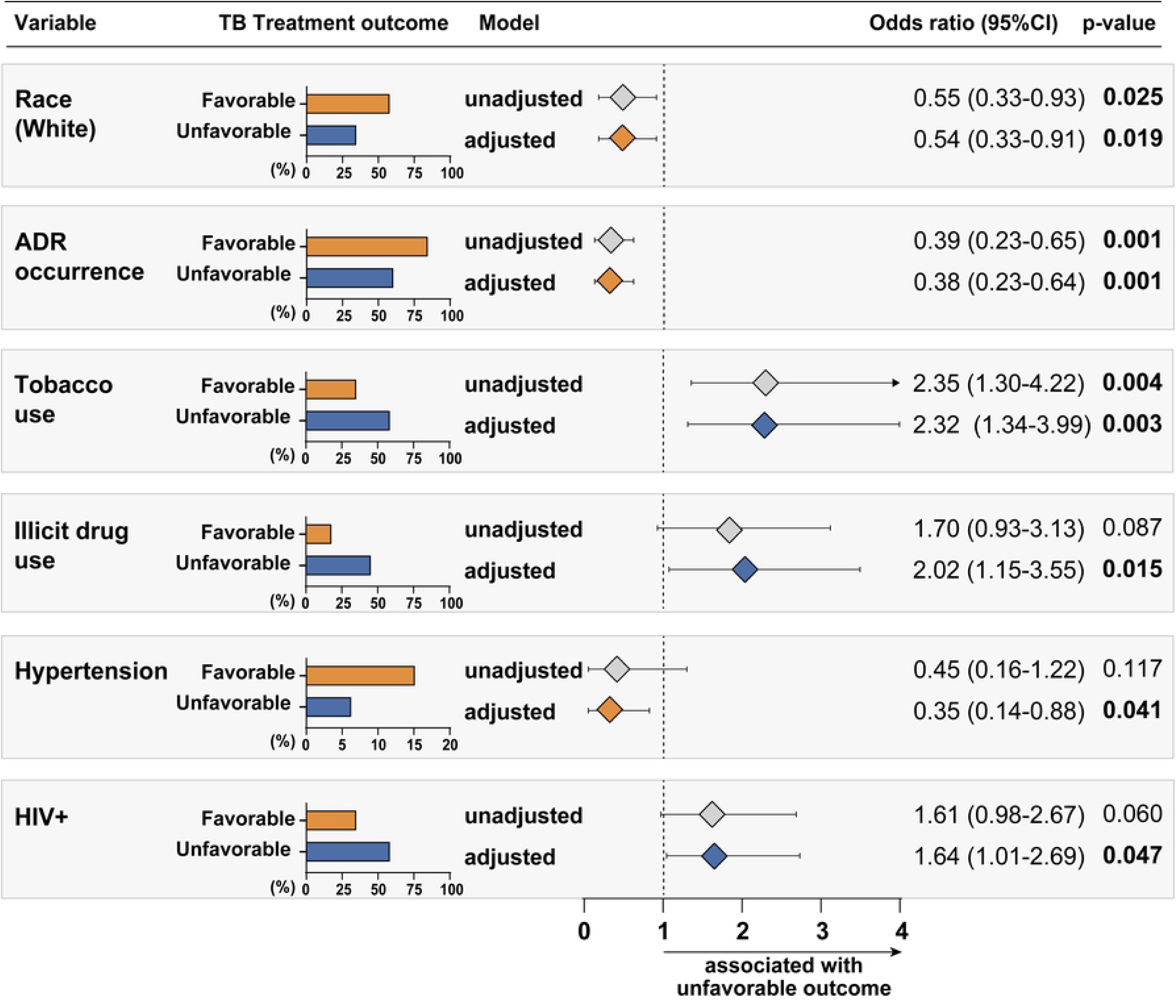
Association between clinical characteristics and tuberculosis treatment outcomes among tuberculosis patients. The logistic binary regression model (backward stepwise regression) was performed to evaluate the independent associations between clinical characteristics, including adverse drug reaction occurrence, of tuberculosis patients and variables with p-value <0.2 results in the univariate analyses (Table 1) and unfavorable treatment outcome. Abbreviation: ADR: adverse drug reaction; CI: confidence interval; TB: tuberculosis.

We compared the clinical characteristics and ADR presentation of patients according to HIV status and noted that there were significant differences between the groups **(Table 3)**. Self-reported race, marital status, smoking habits, alcohol abuse, illicit drug use in the PLHIV group were associated with unfavorable outcomes. ADR occurred in 78.6% of total participants and 72,8% of them were in the PLHIV. Although diagnosis of viral hepatitis serology was included in our model, it was not associated with unfavorable outcomes, but the number of positive cases was small and we did not test viral load to confirm active hepatitis. The main finding of this investigation was that PLHIV experienced more severe forms of ADR than HIV-unexposed participants, especially liver and biliary system disorders **(Table 4)**.

**Table 3.**
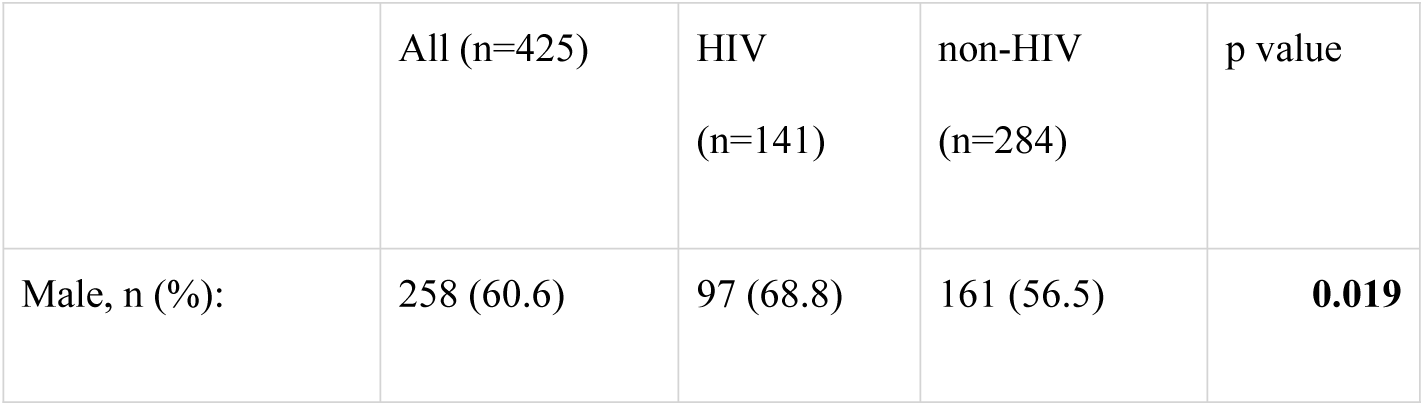
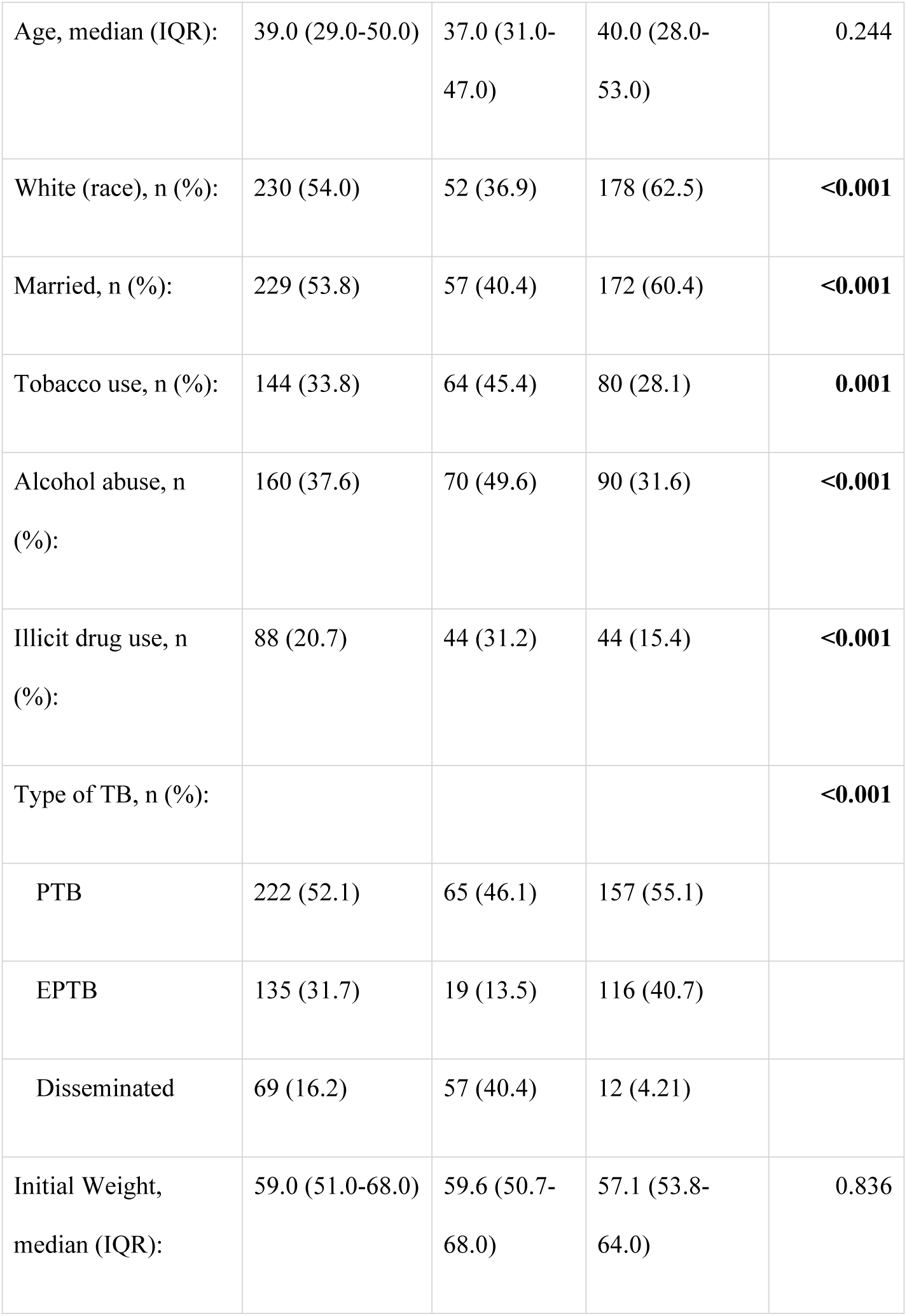
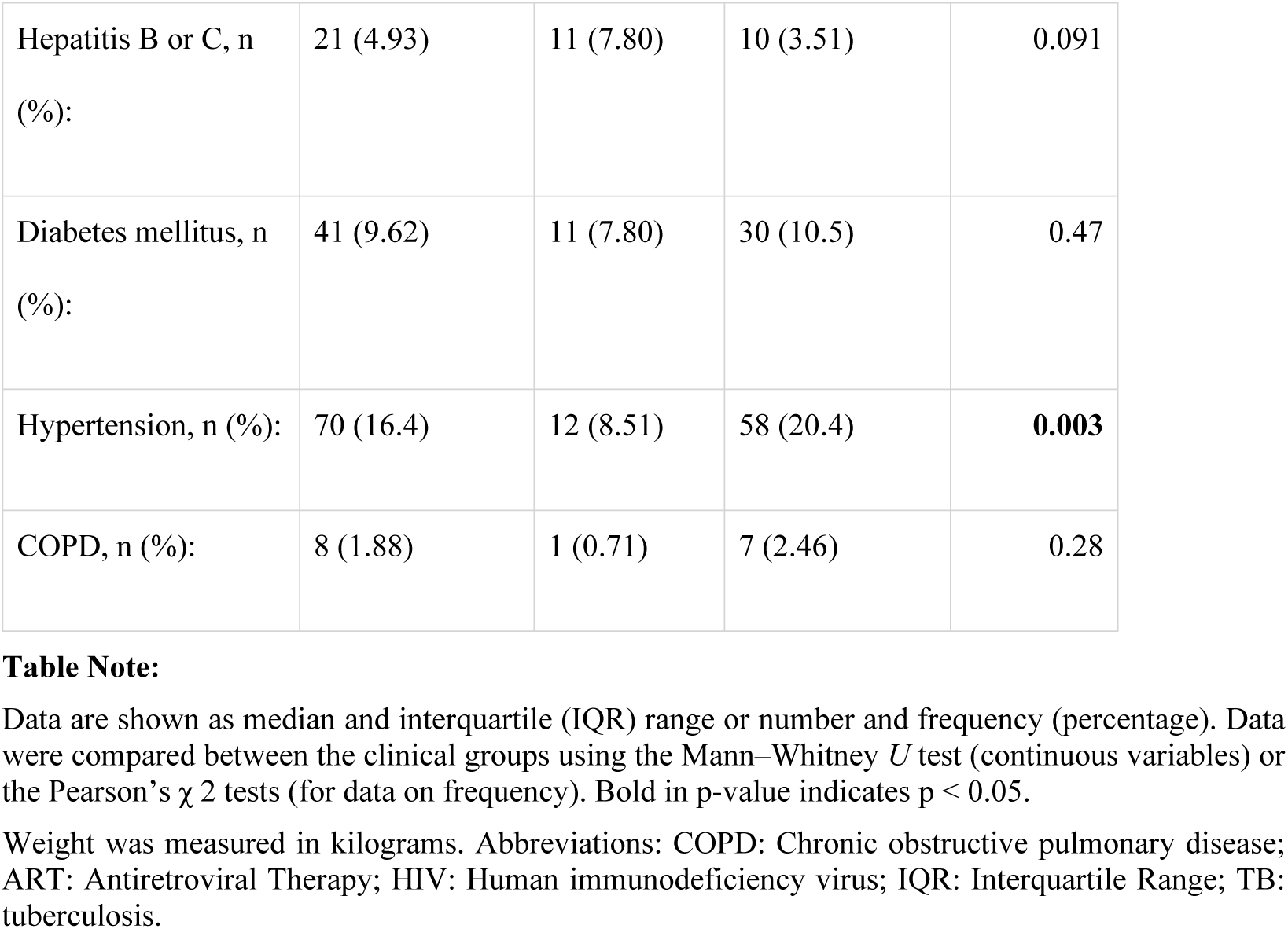
Sociodemographics and clinical characteristics of patients according to HIV status.

|  | All (n=425) | HIV<br>(n=141) | non-HIV<br>(n=284) | p value |
| --- | --- | --- | --- | --- |
| Male, n (%): | 258 (60.6) | 97 (68.8) | 161 (56.5) | **0.019** |
| Age, median (IQR): | 39.0 (29.0-50.0) | 37.0 (31.0-47.0) | 40.0 (28.0-53.0) | 0.244 |
| White (race), n (%): | 230 (54.0) | 52 (36.9) | 178 (62.5) | **<0.001** |
| Married, n (%): | 229 (53.8) | 57 (40.4) | 172 (60.4) | **<0.001** |
| Tobacco use, n (%): | 144 (33.8) | 64 (45.4) | 80 (28.1) | **0.001** |
| Alcohol abuse, n (%): | 160 (37.6) | 70 (49.6) | 90 (31.6) | **<0.001** |
| Illicit drug use, n (%): | 88 (20.7) | 44 (31.2) | 44 (15.4) | **<0.001** |
| Type of TB, n (%): |  |  |  | **<0.001** |
| PTB | 222 (52.1) | 65 (46.1) | 157 (55.1) |  |
| EPTB | 135 (31.7) | 19 (13.5) | 116 (40.7) |  |
| Disseminated | 69 (16.2) | 57 (40.4) | 12 (4.21) |  |
| Initial Weight, median (IQR): | 59.0 (51.0-68.0) | 59.6 (50.7-68.0) | 57.1 (53.8-64.0) | 0.836 |
| Hepatitis B or C, n (%) | 21 (4.93) | 11 (7.80) | 10 (3.51) | 0.091 |
| Diabetes mellitus, n (%) | 41 (9.62) | 11 (7.80) | 30 (10.5) | 0.47 |
| Hypertension, n (%) | 70 (16.4) | 12 (8.51) | 58 (20.4) | **0.003** |
| COPD, n (%) | 8 (1.88) | 1 (0.71) | 7 (2.46) | 0.28 |

**Table 4.**
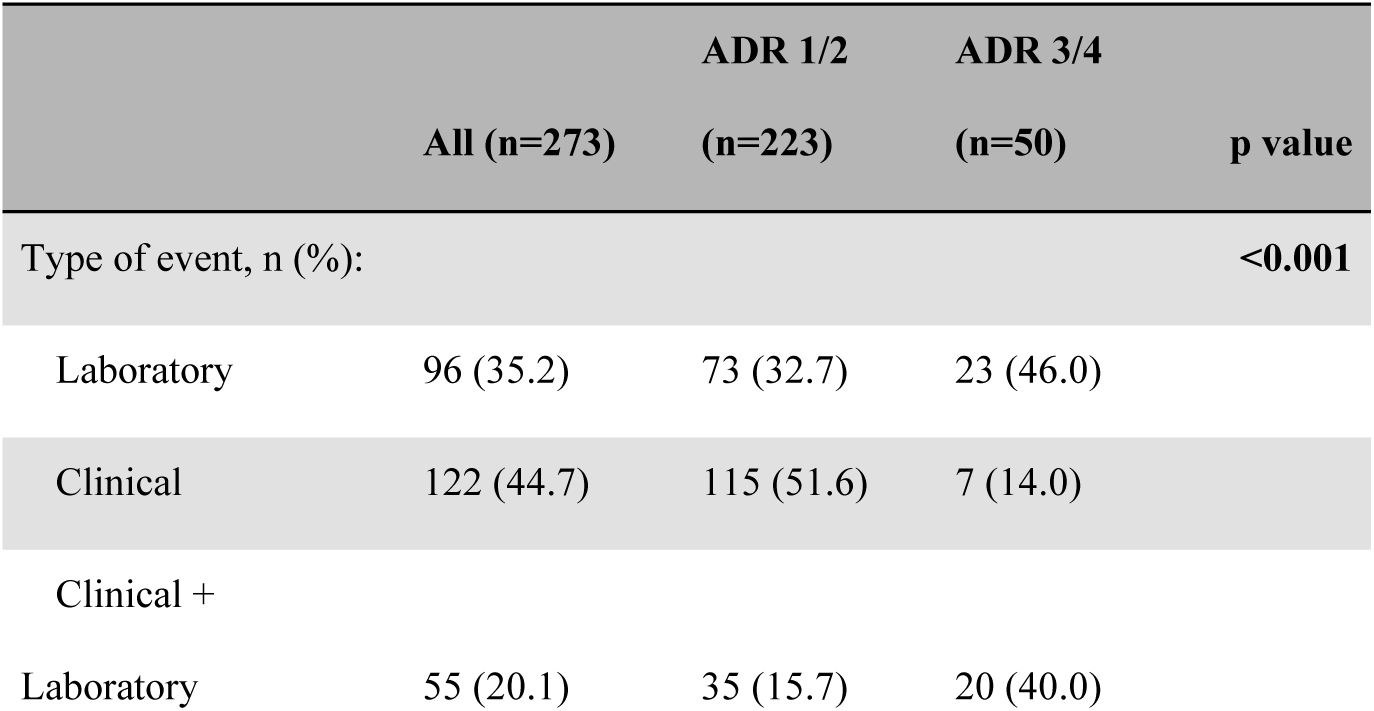
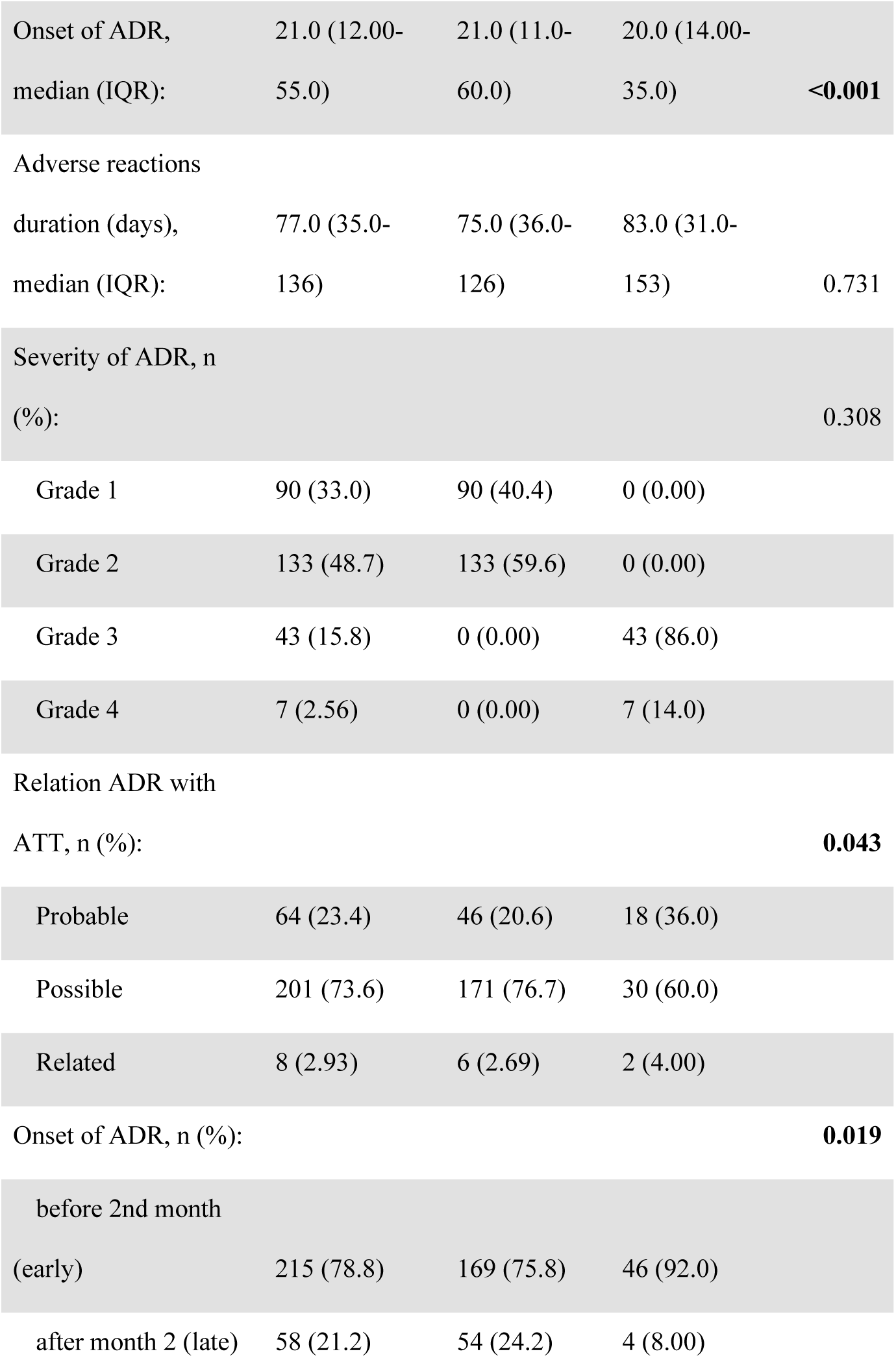
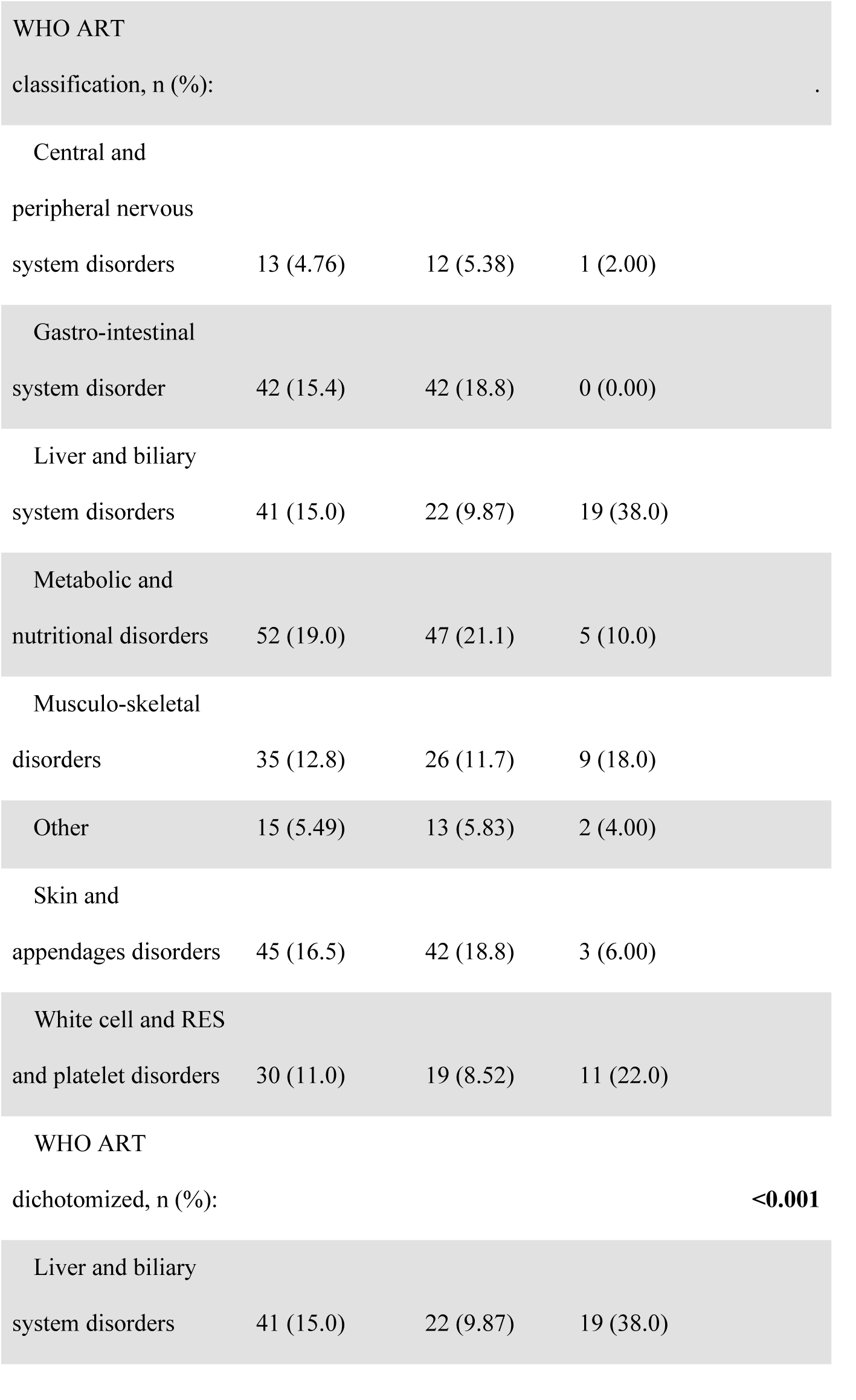
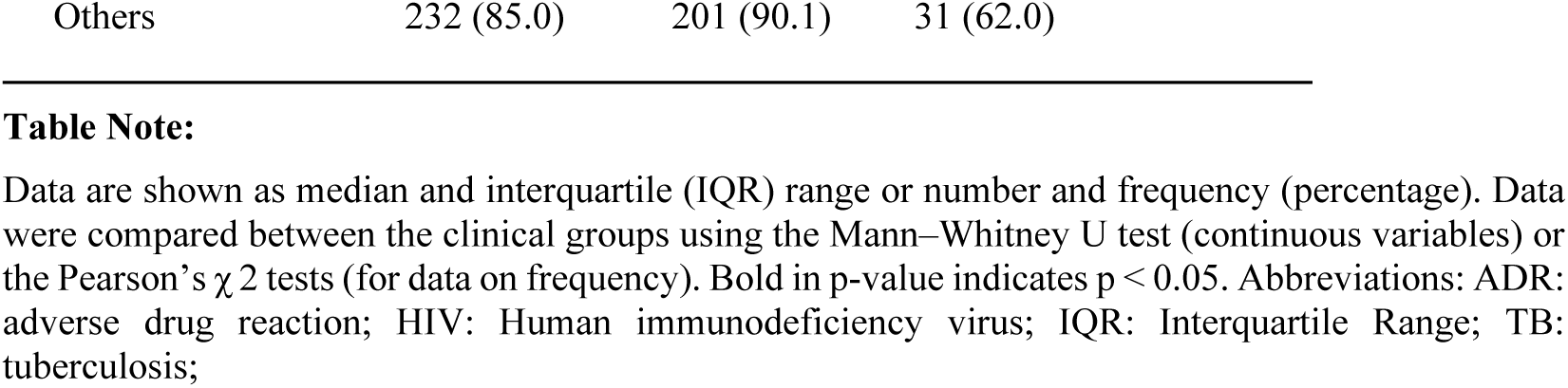
ADR characteristics according to intensity of ADR in PLHIV.

According to clinical data, it was observed that among PLHIV, median CD4 count was 166 cell/μl and those with ADR grade 3 and 4 had lower CD4 count compared to those with ADR grade 1 and 2 and most of those with ADR grade 3/4 displayed CD4 count below 100/uL **(Table 5)**. The characteristics of ADR differed between cases of grade 1/2 compared to those graded 3/4 in terms of type, time of onset, relationship with treatment, and affected system. The most affected system in patients with grade 3/4 ADR was the liver and biliary system **(Table 5)**. Considering all PLHIV, 53.1% experienced unfavorable outcomes (p<0.001). Among them, 183 (33.3%) were ART-experienced while 38.7% ART naïve. In the group of ART experienced patients, 48 (49%) experienced unfavorable outcomes (p<0.001).

**Table 5.** Clinical and laboratory characteristics according to intensity of ADR in PLHIV.

|  | All (n=140) | 1/2 (n=114) | 3/4 (n=26) | p value |
| --- | --- | --- | --- | --- |
| Type of TB, n (%): |  |  |  | **<0.001** |
| PTB | 65 (46.1) | 55 (48.2) | 10 (38.5) |  |
| EPTB | 19 (13.5) | 16 (14.0) | 3 (11.5) |  |
| Disseminated | 57 (40.4) | 43 (37.7) | 13 (50.0) |  |
| HIV Viral Load,<br>median (IQR): | 4.53 (3.26-5.33) | 4.44 (3.21-5.40) | 4.86 (3.55-5.21) | 0.871 |
| CD4 (cells/uL),<br>median (IQR): | 166 (43.0-353) | 176 (52.8-360) | 110 (26.8-180) | 0.065 |
| CD4 levels, n (%): |  |  |  | **0.033** |
| <100 | 56 (40.0) | 43 (37.8) | 13 (50.0) |  |
| 100-200 | 30 (21.4) | 21 (18.4) | 9 (34.6) |  |
| >200 | 54 (38.6) | 50 (43.9) | 4 (15.4) |  |
| Initial Weight,<br>median (IQR): | 59.0 (51.0-68.0) | 59.6 (50.7-68.0) | 57.1 (53.8-64.0) | 0.836 |
| Hepatitis B or C, n<br>(%): | 11 (7.86) | 11 (9.65) | 0 (0.0) | 0.091 |
| Diabetes mellitus, n<br>(%): | 10 (7.14) | 7 (6.1) | 3 (11.5) | 0.47 |
| Hypertension, n (%): | 10 (7.14) | 8 (7.01) | 2 (7.7) | 1.000 |
| COPD, n (%): | 1 (0.71) | 1 (0.88) | 0 (0.0) | 1.000 |

## Discussion

The impact of unsuccessful ATT can directly impair the strategies for TB control [22,23]. The main findings of our study were: ADR was not associated with unfavorable outcomes and PLHIV had higher ADR intensity than HIV-unexposed study participants, which were mainly liver and biliary system disorders, especially those with lower CD4 counts.

As expected, most patients who were lost to follow up had licit or illicit drug addiction. These findings are of great importance for planning strategies to decrease harm but increase early ART, even with the risk of drug to drug interaction. As there was a considerable LTFU, representing the predominant unfavorable outcome, it is urgent to create policies to increase treatment adherence.

In our data male sex was found to be significantly associated with unfavorable outcomes. A possible explanation is that men were less likely to regularly follow treatment when compared with women in our setting [15]. In our study, low schooling, smoke habit and illicit drug use were associated with unfavorable outcomes. However, among PLHIV, alcohol abuse was the only independent factor associated with unfavorable outcomes. Other studies in the literature have shown the same correlation between alcohol abuse [2,24,25] and in the general population, smoke habit was associated with unfavorable outcomes as well as in our study [26].

ADRs were not associated with unfavorable outcomes, contrary to our primary expectations. Probably, patients who experienced ADR were better managed and/or more closely monitored, which contributed to feeling well cared for and having a good adherence to therapy. In contrast, a previous prospective study in Rwanda with TB patients, with or without HIV-infection, has shown that ADRs were associated with an almost two-fold increased risk of LTFU [2].

Another correlation observed in our study was that high schooling participants did not present any unfavorable outcome, which was not a surprise since they probably have a better understanding about the importance of treatment adherence for TB cure. Definitive treatment interruption before cure can lead to recurrence, development of drug resistance, increased costs and a worse epidemiologic situation [27].

We also explored the liver and biliary system disorders in advanced HIV patients and a high incidence of hepatotoxicity was associated with low CD4 count, mainly in those with less than 100 cells//uL. This important finding should help the management of TB in patients with HIV, since more attention will be directed to this type of ADR in this population. Most ART naïve patients are diagnosed with HIV due to TB, as HIV serology is done in all patients with TB diagnosis [11]. These are, in general, the most severe cases and hospitalization is sometimes necessary not only for TB diagnosis but also to manage severe and serious ADR after ATT introduction. The early ART start in these cases is of special interest due to the added risk of hepatotoxicity. Sometimes, when ADR occurs, it is necessary to stop all drugs and reintroduce one by one to identify the causality. Many studies also have found that PLHIV using concomitant ART and ATT had a higher frequency of ADR and low adherence to treatment [27– 29].

In our study we found 26% of PLHIV developing unfavorable outcomes. ART-naïve patients presented a higher incidence of ADR in comparison with ART-experienced PLHIV, although the latter group more frequently developed unfavorable outcomes, especially LTFU. Most of these patients were already cared for at the study referral center or other health clinics and had TB due to low adherence to ART. These patients are different from ART-naïve patients that had a recent TB diagnosis and were tested for HIV for the first time.

In our routine care, ART commencement occurs as soon as possible in the first month of TB treatment for those with low CD4 counts, as well as cotrimoxazole prophylaxis, since this approach has already been proved to reduce mortality [30]. Although all those challenges reported here, our TB lethality was relatively low in PLHIV which shows that the strategy of early ART start works and ADR can be successfully managed.

In conclusion male sex, low schooling, smoke habit, and illicit drug use were all independently associated with unfavorable outcomes during ATT. In PLHIV, alcohol abuse and previous ART use were factors associated with unfavorable outcomes. ADR was more frequent in the beginning of TB treatment and was not associated with unfavorable outcomes.

In our study, clinical center patients were attended by different physicians who in general conduct the whole treatment and have laboratory exams available to help ADRs detection and management. Also, medical care was available at the hospital every day, 24 hours a day, which helps ADR management any time they occur. We had some limitations: directly observed treatment (DOT) was not performed in our site due to our characteristics of being a tertiary hospital and far from most patients’ houses. Viral hepatitis were screened with serology and no viral load was performed, the low proportion of viral hepatitis in our study limited the analyses in this group.

The identification of factors associated with unfavorable outcomes could allow health care to appropriately risk-stratify patients for closer management and improve outcomes. In addition, investments in programs destined to reduce tobacco, alcohol and drug addiction are also necessary to improve TB treatment adherence and decrease LTFU. On the other hand, an investment in education is necessary by the government to make patients better understand the disease and the importance of treatment completion.

## Data Availability

All relevant data are within the manuscript and its Supporting Information files.

## Author Contributions

FS, CS and VR contributed to conception and design of the study. FS collected the data and organized the initial database. MA-P and MA performed the statistical analysis and data visualization. VR and BA supervised the project execution. All authors contributed to manuscript writing, revision, read, and approved the submitted version.

## Acknowledgments

The authors thank the study participants.

## Supplementary material

**S1 File**. Raw data.

